# A comparative analysis of the performance of chatGPT4, Gemini and Claude for the Polish Medical Final Diploma Exam and Medical-Dental Verification Exam

**DOI:** 10.1101/2024.07.29.24311077

**Authors:** Dorota Wójcik, Ola Adamiak, Gabriela Czerepak, Oskar Tokarczuk, Leszek Szalewski

## Abstract

In the realm of medical education, the utility of chatbots is being explored with growing interest. One pertinent area of investigation is the performance of these models on standardized medical examinations, which are crucial for certifying the knowledge and readiness of healthcare professionals. In Poland, dental and medical students have to pass crucial exams known as LDEK (Medical-Dental Final Examination) and LEK (Medical Final Examination) exams respectively. The primary objective of this study was to conduct a comparative analysis of chatbots: ChatGPT-4, Gemini and Claude to evaluate their accuracy in answering exam questions of the LDEK and the Medical-Dental Verification Examination (LDEW), using queries in both English and Polish. The analysis of Model 2, which compared chatbots within question groups, showed that the chatbot Claude achieved the highest probability of accuracy for all question groups except the area of prosthetic dentistry compared to ChatGPT-4 and Gemini. In addition, the probability of a correct answer to questions in the field of integrated medicine is higher than in the field of dentistry for all chatbots in both prompt languages. Our results demonstrate that Claude achieved the highest accuracy in all areas analysed and outperformed other chatbots. This suggests that Claude has significant potential to support the medical education of dental students. This study showed that the performance of chatbots varied depending on the prompt language and the specific field. This highlights the importance of considering language and specialty when selecting a chatbot for educational purposes.

## Introduction

Artificial intelligence (AI) is a transformative technology with the potential to revolutionize various aspects of our lives by enhancing efficiency, enabling new capabilities, and providing personalized experiences. Various applications are available, such as LLM, generative AI. Generative AI refers to a subset of artificial intelligence focused on creating new content or data that is similar to existing data. It uses machine learning models to generate text, images, music, and other media. A Large Language Model is a type of AI model specifically designed to understand, generate, and manipulate human language on a large scale. LLMs are built using deep learning techniques and trained on vast amounts of text data to generate human-like text. These models leverage deep learning techniques, specifically the Transformer architecture, to process and produce coherent, contextually relevant responses to a wide array of queries ^1,2,3^.

In the realm of medical education, the utility of chatbots is being explored with growing interest. One pertinent area of investigation is the performance of these models on standardized medical examinations, which are crucial for certifying the knowledge and readiness of healthcare professionals.

In Poland, dental and medical students have to pass crucial exams known as LDEK(Medical-Dental Final Examination) and LEK (Medical Final Examination) exams respectively. These exams are crucial in determining whether a student readiness to enter the medical profession. Evaluating the performance of AI chatbots on such examinations could provide valuable insights into how artificial intelligence might support medical education. The LEK, which is comparable to the United States Medical Licensing Examination (USMLE), is particularly significant. After passing this exam, candidates are eligible to apply for a medical licence in Poland. In addition, this qualification is recognised in all EU Member States due to European Union regulations outlined in Directive 2005/36/EC. This qualification is recognized across EU member states, allowing for potential practice throughout the Union.^4^

This paper aims to assess the performance of ChatGPT4, Claude and Gemini (chatbots) on the LDEK, examining its accuracy, strengths, and limitations in this context. Through this investigation, we seek to understand the potential role of chatbots in medical education and their implications for the future of healthcare training and practice.

## Materials and Methods

### Study design

The primary objective of this study was to conduct a comparative analysis of chatbots: ChatGPT-4, Gemini and Claude to evaluate their accuracy in answering exam questions of the LDEK and the Medical-Dental Verification Examination (LDEW), using queries in both English and Polish. Study design is presented in the flowchart (Figure 1).

Several secondary objectives were also formulated.

The first secondary objective was to determine the relationship between the self-assessed confidence of the chatbots and the percentage of correct answers in the population as reported by the Medical Examination Centre (CEM).

The second secondary objective was to determine whether the self-assessed confidence was related to the actual accuracy of the chatbots.

The third secondary objective was to investigate and evaluate the agreement of the responses generated by each chatbot between prompts in English and Polish languages.

The fourth subsidiary objective involved evaluating the agreement of responses across three measurements within each prompt language and among the chatbots.

The fifth subsidiary objective was to ascertain if significant differences in accuracy existed among the chatbots within identical prompt languages.

The sixth subsidiary objective was to rigorously assess the existence of disparities in accuracy between the prompt languages employed by the same chatbot.

The final, seventh subsidiary objective was to conduct a detailed evaluation of the variations in accuracy among chatbots across different categories of questions (specifically, those pertaining to the seven distinct disciplines within the dental faculty: conservative dentistry, paediatric dentistry, dental surgery, prosthetics, periodontology, orthodontics, and integrated medicine) and between various prompt languages.

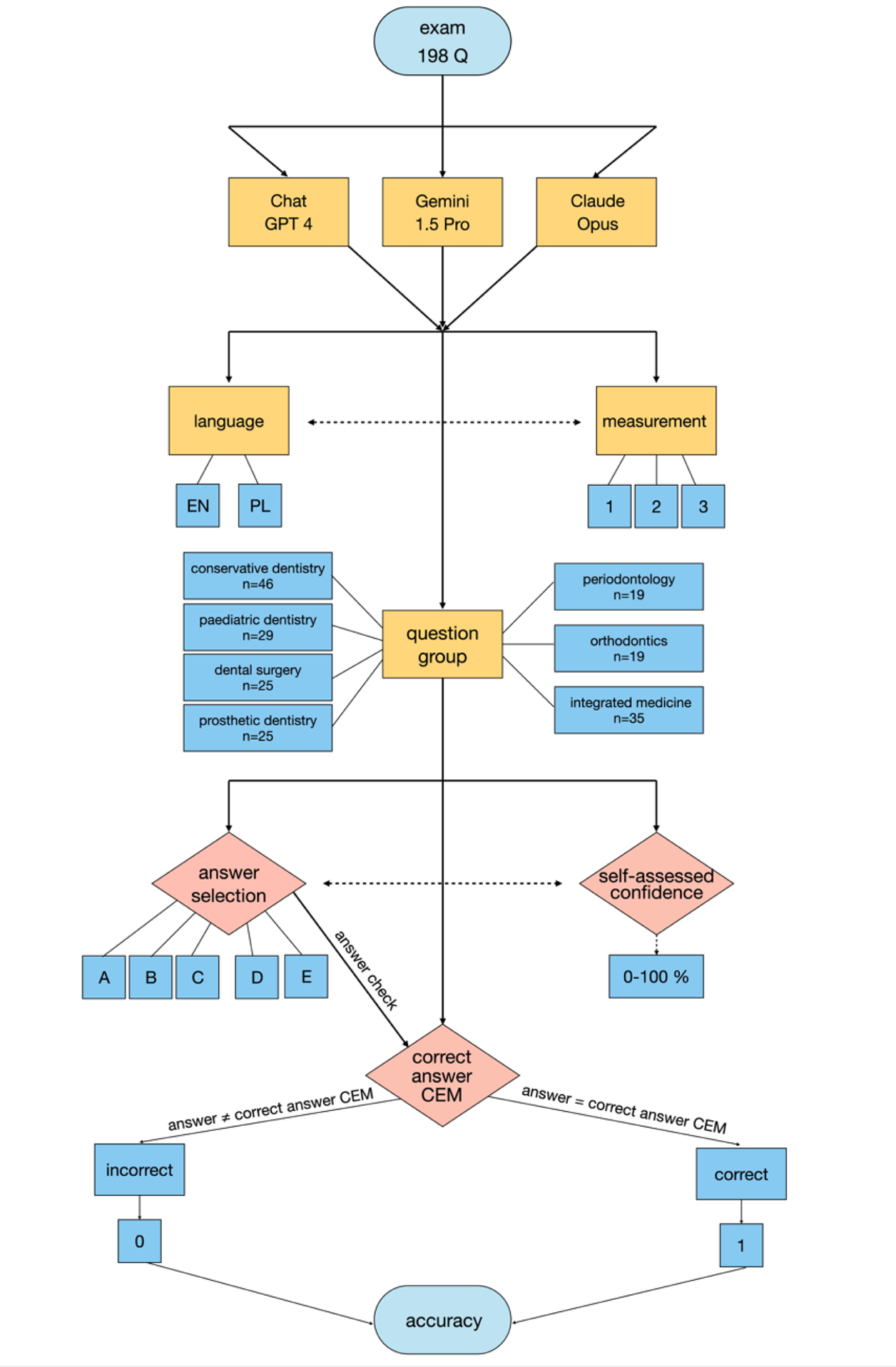

#### Questions

The study was conducted from 27 March to 13 April. Questions from the LDEK and LDEW exams conducted in April 2024 were used for the study. These exams, provided by the Central Examination Board (CEM), comprised 198 questions each. The LDEK exam was held in Polish, while the LDEW exam was in English. Both exams contained identical questions, differing only in the language of presentation. Each question was preceded by a pre-standardized prompt in Polish or English, depending on the exam version. Detailed information, including the content of a sample prompt and the exam questions, can be found in the Appendix.

#### Question group

The study examined responses to questions from seven areas of dentistry and medicine (see Table 1 for a detailed description of the variables). For the purpose of this study, the question groups emergency medicine, bioethics and medical law, medical certification and public health were combined into one category – integrated medicine. Each question was a multiple-choice question with five possible answers from A to E.

#### Languages and measurements

For the data collection phase, the official websites of the chatbot providers were utilized: responses from ChatGPT were sourced from https://chatgpt.com, those from Gemini were obtained from https://deepmind.google/technologies/gemini/, and responses from Claude were accessed via https://console.anthropic.com. A single prompt was used to elicit responses to one question, employing a total of 198 prompts per examination. To ensure the internal consistency of the responses, the examination was repeated three times with prompts in Polish and three times with prompts in English, with intervals of a few hours between each session. In total, information from 1,188 prompts per chatbot was collected.

#### Answer selection and self-assessed confidence

The chatbot was asked to select a correct answer and provide a self-assessed confidence, which represents the chatbot’s own assessment of the probability that its answer to the test question was correct. The self-assessed confidence was to be expressed on a scale of 0% to 100%, with 0% representing complete uncertainty about the accuracy of the answer and 100% representing absolute confidence in the accuracy of the answer.

The study involved the use of a rather complex prompt, which required chatbots to apply many skills that went beyond simple verbal reasoning. Each chatbot had to use its skills to answer single-choice questions and estimate the probability of each option being correct. To answer single-choice questions, the chatbots had to demonstrate comprehension skills, i.e. understand the content of the question and the possible answers. Knowledge retrieval was important in order to find relevant information from the internal knowledge database. Decision-making involved selecting the most likely answer from the available options, while pattern recognition enabled the chatbots to recognise patterns in the questions and answers, which was crucial for making the right choice.

Furthermore, to estimate the probability of each option being correct, the chatbots had to apply probabilistic reasoning, which enabled them to assess the likelihood of each answer being correct. The confidence assessment involved evaluating their own certainty regarding the chosen answer. Statistical analysis was employed to analyse the data and evaluate the answers through statistical methods. Risk assessment enabled the chatbots to evaluate the risk of an incorrect answer and its potential consequences.

#### Phase of answer verification

The correctness of the response provided by the chatbot to the examination question, which involved selecting the letter corresponding to the answer option, was subsequently verified using a key provided by CEM. If the response matched the key, it was considered correct; otherwise, it was deemed incorrect.

### Hypotheses

The investigation is segmented into several key hypotheses, each aiming to dissect distinct aspects of chatbot performance. This includes examining the correlation between self-assessed and actual accuracy, the consistency of responses across different languages, and the comparative performance of different chatbot models. Additionally, the research extends to evaluating the performance of chatbots on specific domains of knowledge and across different types of queries, thereby providing a comprehensive overview of their capabilities and limitations. For a comprehensive list of hypotheses, please refer to Appendix A.

### Statistical analysis

The significance level of the statistical tests was set at α = 0.05. The distribution of variables was described using descriptive statistics. Measures for the central tendency, in particular the mean (*M*) and the variance in the form of the standard deviation (SD), were used. Categorical variables were described using frequencies across individual categories (*n*) and percentages.

#### Correlation analysis

The association between a dichotomous variable and a numeric variable was assessed using the rank biserial correlation coefficient (^r^^ biserial rank). Spearman’s rank correlation coefficient (rho) was used to measure the strength and direction of the association between two numerical variables, especially for non-normally distributed variables. The rank biserial coefficient was interpreted on the basis of Funder convention (Funder, 2019)^5^. The rho coefficient was interpreted on the basis of the Cohen convention (Cohen, 1988)^6^.

#### Analysis of the agreement

For each of the three chatbots under investigation, the linguistic consistency between their Polish and English versions of answer was evaluated using Cohen’s Kappa coefficient. The results of Cohen’s Kappa were interpreted in accordance with McHugh’s guidelines (McHugh, 2012)^7^.

The agreement between the three responses of the same chatbot to the same exam question, evaluated separately in both Polish and English, was calculated using Fleiss’ Kappa coefficient. Each response was treated as an independent rater for the purpose of this evaluation. The results were interpreted on the basis of Fleiss’ convention (Fleiss, 2003)^8^.

### Multivariate analysis

#### Generalised linear mixed model (GLMM)

Generalised linear mixed models (GLMM) were used to assess the probability of correct responses and to enable comparisons between specific chatbots as well as to estimate effect sizes.

The choice of this methodology was motivated by several factors. A multivariate approach was required due to the complexity of the data, which included multiple variables affecting the accuracy of chatbot responses. A generalised model was chosen due to the dichotomous nature of the outcome variables (accuracy), which violates the assumptions of classical linear regression. The implementation of a mixed model was essential to account for repeated measurements, as each chatbot provided three responses to the same exam question.

Two GLMM models were implemented:

Model 1 aimed to predict and determine the effects of chatbot type and query language on response accuracy.

Model 2 was developed to determine the influence of question group, chatbot type and language on the probability of a correct answer.

In both models, the dichotomous outcome variable was fitted using a binomial distribution with a logit link function. Detailed model specifications can be found in Appendix X.

In addition, the use of GLMM enables the conduct of contrast analysis, allowing for more nuanced comparisons between specific groups or experimental conditions. Detailed specifications of Model 1 and Model 2 can be found in Appendix.

#### Specification of fixed and random effects in GLMER

Two Generalized Linear Mixed-Effects Models (GLMER) were implemented to account for dependencies between observations and to analyze data with repeated measurements.

Model 1: accuracy_chat ~ chatbot * language + (1|ID_Q)

This model examined the interaction between chatbot type and prompt language on response accuracy. Fixed effects included chatbot type, language, and their interaction. The random effect was the random intercept for questions (ID_Q).

Random effects for Model 1 were quantified using:

- σ^2^: Residual variance
- τ00: Variance of the random intercept for questions
- ICC: Intraclass Correlation Coefficient, indicating the proportion of variance explained by the grouping structure

Model 2: accuracy_chat ~ chatbot * Q_group * language + (language|ID_Q)

This model investigated the interaction between question group, chatbot type, and prompt language on response accuracy. Fixed effects included chatbot type, question group, language, and their interactions. Random effects included random intercepts for questions and random slopes for language by question.

Random effects for Model 2 were quantified using:

- σ^2^: Residual variance
- τ00 ID_Q: Variance for the random intercept for questions
- τ11 ID_Q.language pl: Variance for the random slope of Polish language by question
- ρ01 ID_Q: Correlation between the random intercept for questions and the random slope of Polish language

Both models used a binomial distribution with a logit link function. The models were fitted using maximum likelihood estimation, specifically the Laplace approximation.

#### Target effects and interactions

Model 1 target effect: The interaction between chatbot type (ChatGPT-4, Gemini, Claude) and prompt language (English, Polish), with prompt language as a moderator.

Model 2 target effect: The three-way interaction between question group, chatbot type (ChatGPT-4, Gemini, Claude), and prompt language (English, Polish), with language as a moderator.

Goodness-of-fit for both models was assessed using conditional and marginal coefficients of determination (Marginal R^2^ for fixed effects, Conditional R^2^ for both fixed and random effects).

#### Optimisation and model fitting

For Model 2, optimisation was performed using the Bound Optimisation BY Quadratic Approximation (BOBYQA) algorithm, developed by M.J.D. Powell (2009). This algorithm was chosen to handle the complexity and size of the data involved. Model 1 did not require additional optimisation procedures beyond the standard fitting process^9^.

#### Event and interaction effect

The event was defined as the correct answer to the question asked. The target effect in Model 1 examined the interaction between chatbot type (ChatGPT-4, Gemini, Claude) and prompt language (English, Polish). This model evaluated how the language of the prompt influenced the accuracy of responses provided by each chatbot.

For Model 2, the target effect analyzed the interaction between the question group, the chatbot type (ChatGPT-4, Gemini, Claude), and the prompt language (English, Polish), with the prompt language acting as a moderator. This model explored how different question groups influenced the accuracy of responses from various chatbots in English and Polish within each subgroup.

#### Hypothesis tests and multiple comparisons

To test hypotheses about specific differences between groups, an estimation of marginal means (EMM) and a contrast analysis were performed. The Holm correction was applied to adjust significance levels when comparing three or more groups, controlling the family-wise error rate in cases of multiple pairwise comparisons.

In the EMM analysis, values of the outcome variable in subgroups were presented as probabilities of obtaining a correct response. For the contrast analysis, these values were transformed into odds ratios (OR). Effect sizes were interpreted based on Cohen’s (1988) convention. P-values were calculated as approximations to Wald Z distribution statistics^6^.

The target effect was calculated in two ways: first, analyzing differences between chatbots within language categories, and second, examining differences between languages for each chatbot. Detailed model specifications can be found in the Appendix.

#### Statistical environment

Analyses were conducted using the R Statistical language (version 4.4.0; R Core Team, 2024) on Windows 11 pro 64 bit, using the packages: *readxl*^10^ and *dplyr* ^11^*, effectsize*^12^, *lme4*^13^, *sjPlot*^14^, *irr*^15^, *tidyr*^16^*, ggstatsplot*^17^, *ggplot2*^18^.

### Characteristics of the study sample

The existing dataset comprised 3564 observations and included the following variables: a unique identifier for each exam question, the category of the question, the language of the prompt, which was either Polish or English, the model of the chatbot used, including ChatGPT-4, Gemini and Claude, the identifier of the rater, labelled as A, B or C, the repetition number, which ranged from 1 to 3, the accuracy of the chatbot response, coded as 1 for correct and 0 for incorrect, self-assessed confidence and the percentage of correct answers as reported by the Medical Examination Centre.

The questions were distributed across the individual areas as follows: conservative dentistry with endodontics (23%), pediatric dentistry (15%), oral surgery (13%), prosthetic dentistry (13%), periodontology (10%), orthodontics (10%), and integrated medicine (18%).

Missing data were observed in one variable: self-assessed confidence (n = 1).

### Descriptive statistics

Claude shows the highest mean accuracy in most specialties (Table 1), both in English and Polish. The best results were achieved in the specialty of integrated medicine, where accuracy reached 95% for English and 92% for Polish. ChatGPT-4 and Gemini show more varied results.

The biggest differences between the chatbots were observed in the specialty of oral surgery, where Gemini (23%) performs poorly compared to ChatGPT-4 (38%) and Claude (53%) in Polish. In periodontology, Gemini also achieves significantly lower accuracy in Polish (30%) than ChatGPT-4 (54%) and Claude (54%). In the field of integrated medicine, all chatbots have the highest accuracy.

The standard deviations (SD) vary from 1.4 to 6.3, indicating moderate variability in response accuracy. Lower SD values in the area of integrated medicine, indicate an even lower dispersion around the mean value.

All chatbots rate their answers in the area of integrated medicine as the most accurate (Claude has the highest self-assessment values in most areas). Moderate SD values indicate that the chatbots are quite consistent in their assessment of their own answers within the respective subgroups (Table 2).

**Table 1:** Mean accuracy values of the chatbots between language categories and question specialties (N = 1188).

| question group | language | mean accuracy |  |  |
| --- | --- | --- | --- | --- |
|  |  | chatGPT4<br>M (SD) | Gemini<br>M(SD) | Claude<br>M (SD) |
| conservative dentistry<br>n=46 (23%) | English | 57 (3.5) | 54 (3.1) | 72 (3.4) |
|  | Polish | 61 (3.5) | 44 (3.8) | 72 (3.6) |
| paediatric dentistry<br>n=29 (15%) | English | 49 (4.4) | 55 (4.4) | 68 (4.6) |
|  | Polish | 63 (4.8) | 53 (4.9) | 71 (4.8) |
| dental surgery<br>n=25 (13%) | English | 56 (4.5) | 51 (4.5) | 69 (4.9) |
|  | Polish | 40 (4.9) | 23 (3.9) | 55 (5.1) |
| prosthetic dentistry<br>n=25 (13%) | English | 44 (4.9) | 51 (4.4) | 45 (5.2) |
|  | Polish | 52 (4.9) | 51 (4.3) | 39 (5.1) |
| periodontology<br>n=19 (10%) | English | 49 (5.8) | 54 (5.3) | 60 (5.9) |
|  | Polish | 54 (6.3) | 30 (4.5) | 54 (6.2) |
| orthodontics<br>n=19 (10%) | English | 42 (5.6) | 56 (5.2) | 54 (5.6) |
|  | Polish | 61 (5.5) | 44 (5.2) | 56 (5.4) |
| integrated medicine<br>n=35 (18%) | English | 90 (2.4) | 75 (3.2) | 95 (1.9) |
|  | Polish | 82 (2.7) | 81 (3.2) | 92 (2.4) |

**Table 2:** Mean self-assessed confidence values of the chatbots for different language categories and question specialties (N = 1188).

| question group | language | self-assessed confidence |  |  |
| --- | --- | --- | --- | --- |
|  |  | chatGPT4<br>M (SD) | Gemini<br>M(SD) | Claude<br>M (SD) |
| conservative dentistry<br>n=46 (23%) | English | 62 (2.3) | 66 (2.1) | 78 (1.9) |
|  | Polish | 69 (2.4) | 70 (2.3) | 71 (1.6) |
| paediatric dentistry<br>n=29 (15%) | English | 71 (2.4) | 66 (2.3) | 84 (2.1) |
|  | Polish | 76 (2.5) | 89 (1.6) | 71 (1.6) |
| dental surgery<br>n=25 (13%) | English | 64 (2.4) | 65 (2.8) | 76 (3.0) |
|  | Polish | 71 (3.2) | 69 (3.3) | 81 (2.5) |
| prosthetic dentistry<br>n=25 (13%) | English | 59 (2.6) | 67 (3.0) | 72 (2.8) |
|  | Polish | 72 (3.4) | 67 (3.1) | 73 (1.9) |
| periodontology<br>n=19 (10%) | English | 68 (3.7) | 61 (3.9) | 84 (3.0) |
|  | Polish | 85 (3.7) | 63 (4.2) | 90 (1.8) |
| orthodontics<br>n=19 (10%) | English | 60 (2.7) | 78 (3.7) | 71 (3.4) |
|  | Polish | 70 (3.4) | 62 (4.9) | 72 (3.2) |
| integrated medicine<br>n=35 (18%) | English | 78 (2.0) | 75 (1.6) | 90 (1.4) |
|  | Polish | 85 (1.9) | 72 (2.1) | 84 (1.4) |

### Analysis of the percentage of correct answers by language group

The accuracy metric was determined by calculating the arithmetic mean of the three measurements. Calculations were performed separately for each language group to account for possible differences in the quality of the answers given by the chatbots depending on the language of the prompt (Figure 1).

**Figure 1:**
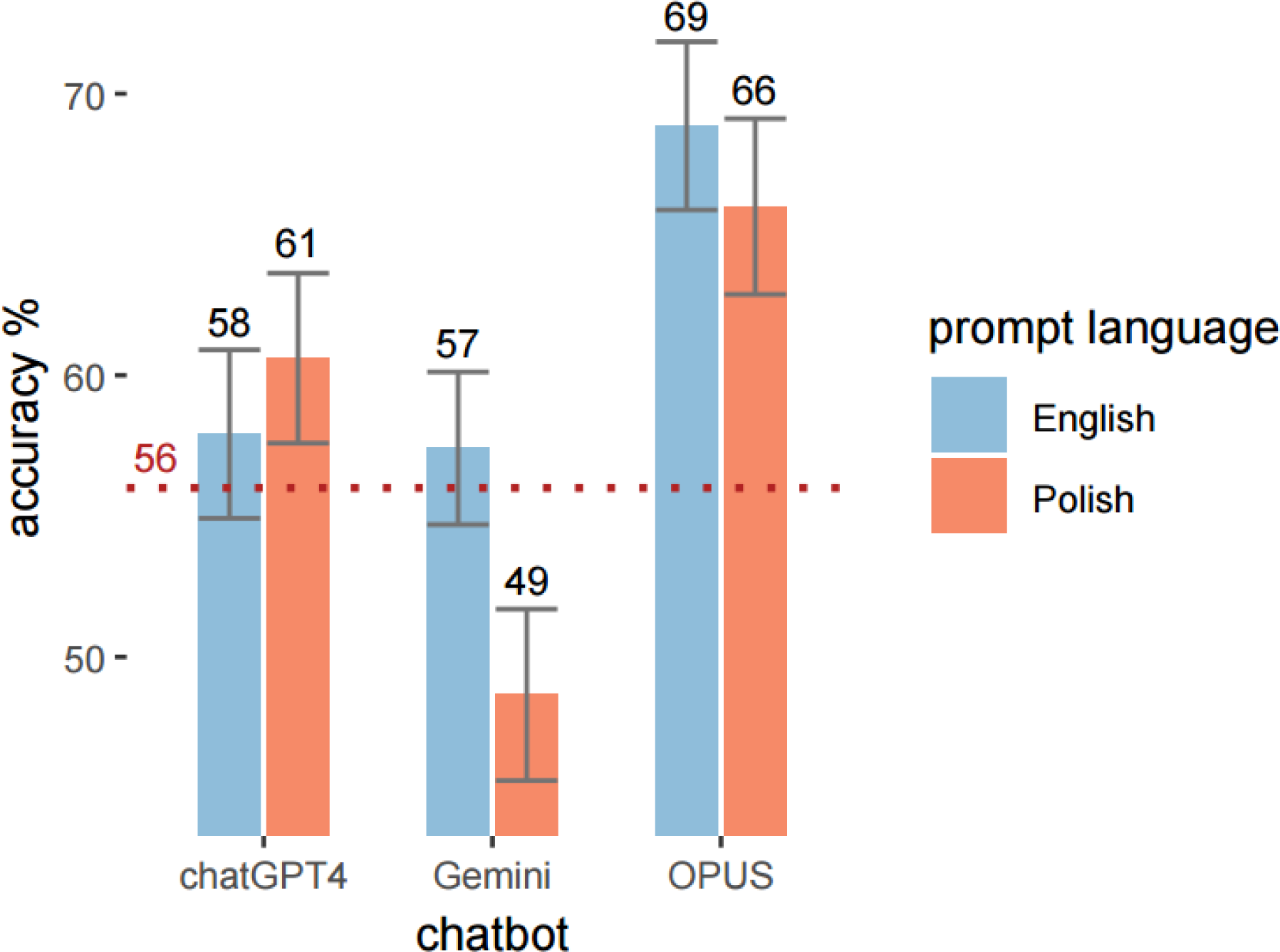
Accuracy by chatbot and language*. *The line at 56% indicates the minimum score for passing the LDEK and LDEW exams

Based on our results, considering the average of three measurements of chatbots answering exam questions, only Claude would pass the exam in each attempt. On the other hand, only Gemini, when taking the exam in Polish, would certainly fail.

## Results

### Correlation analysis between self-assessed confidence and aggregate CEM system test scores for the population

The results of the Spearman correlation analysis (Table 3) show a statistically significant, small, positive correlation between the aggregated CEM system exam results for the population and the self-assessed confidence for ChatGPT-4 and Gemini. The effect size for ChatGPT-4 and Gemini is small.

There is a weak, positive correlation between the self-assessed confidence of ChatGPT-4 and Gemini and the effectiveness of the students’ answers to the exam questions. The more difficult the question is for the students, the lower the chatbot rates its probability of giving a correct answer. The small effect size suggests that the chatbots’ self-assessment may not be a completely reliable indicator of their actual effectiveness. Based on the results of these analyses, the first hypothesis was accepted.

**Table 3:** Spearman correlation results: Self-assessed confidence vs. aggregated CEM test results (N = 1188).

| chatbot | rho | p |
| --- | --- | --- |
| chatGPT4 | 0.16 | < **0.001** |
| Gemini | 0.10 | < **0.001** |
| Claude | 0.04 | 0.192 |

### Correlation between self-assessed confidence and actual accuracy when answering exam questions

There is a statistically significant correlation between self-assessed confidence and the actual accuracy of ChatGPT-4 and Claude in providing correct answers to exam questions (Table 4). The higher the self-assessed confidence, the higher the actual accuracy in answering exam questions correctly. The rank biserial correlation coefficients for ChatGPT-4 and Claude indicate a medium and very small effect size respectively.

There is a positive correlation between the self-assessed confidence of ChatGPT-4 and Claude and their actual effectiveness in answering exam questions. Based on the results of these analyses, the second hypothesis was accepted.

**Table 4:** Rank biserial correlation: self-assessed confidence vs. actual accuracy.

| chatbot<br>(N=1188) | $r_{\text{biserial}}^{\text{rank}}$ | p |
| --- | --- | --- |
| chatGPT4 | 0.22 | < **0.001** |
| Gemini | 0.02 | 0.590 |
| Claude | 0.09 | < **0.001** |

### Analysis of the agreement of chatbot answers to exam questions based on the language of the prompt

In the study, the agreement of answers given by chatbots to exam questions in Polish and English was assessed using Cohen’s Kappa coefficient as a measure of reliability.

**Table 5:** Analysing the agreement of answers to exam questions within the chatbots between the query languages.

| chatbot<br>(N=1188) | Cohen's $\kappa$ | z | p |
| --- | --- | --- | --- |
| Chat GPT4 | 0.45 | 10.9 | < **0.001** |
| Gemini | 0.39 | 9.47 | < **0.001** |
| Claude | 0.59 | 14.3 | < **0.001** |
| Cohen's $\kappa$ - Cohen's Kappa coefficient, z – z-score, | | | |

| p - p value |
| --- |

The Cohen’s Kappa coefficient (Table 5) shows that ChatGPT-4 and Claude indicate moderate agreement between the responses in English and Polish. Gemini shows weak agreement between the responses in English and Polish. All values determined are statistically significant (p < 0.001).

All chatbots analysed differed in the consistency of responses between Polish and English prompts. While there is some agreement between languages for all chatbots, the level of consistency differs. As a result of these analyses, the fourth hypothesis was accepted.

**Table 6:** Analysis of the agreement of answers to exam questions across three measurements, depending on chatbot and prompt language.

| chatbot<br>(N = 594) | prompt language | Fleiss' $\kappa$ | z | p |
| --- | --- | --- | --- | --- |
| Chat GPT4 | Polish | 0.58 | 14.1 | **< 0.001** |
|  | English | 0.59 | 14.3 | **< 0.001** |
| Gemini Gemini | Polish | 0.60 | 14.7 | **< 0.001** |
|  | English | 0.40 | 9.82 | **< 0.001** |
| Claude Claude | Polish | 0.78 | 19.1 | **< 0.001** |
|  | English | 0.72 | 17.6 | **< 0.001** |
| Fleiss' $\kappa$ - Fleiss' Kappa coefficient, z – z-score, p - p value | | | | |

Analysis of the agreement of the three chatbot answers to the same questions using Fleiss’ kappa coefficient (Table 6) revealed differences depending on the chatbot model and the prompt language. The best results, indicating excellent agreement between consecutive measurements, were obtained for Claude chatbot in Polish. ChatGPT-4 also achieved good agreement between measurements, regardless of the language of the responses. Gemini shows the weakest response consistency in English.

All analysed chatbots differ in the degree of response consistency across three measurements in both prompt languages. Chatbot responses to the same questions may vary depending on the subsequent measurement for both languages. The findings from these analyses led to the acceptance of the fourth hypothesis.

### Model 1 fit results

The model was fitted on 3564 observations and 3 raters.

Marginal R^2^ / Conditional R^2^: 0.027 / 0.624

Random effects: σ^2^=3.29, τ00 ID_Q=5.22, ICC=0.61

The model was fitted on 3564 observations. The model explains 2.7% of the variance in the dependent variable when only fixed effects are considered (marginal R^2^ = 0.027), and 62,4% of the variance when both fixed and random effects are considered (conditional R^2^ = 0.624). This means that while the independent variables alone explain only a relatively small part of the variability in the responses, the inclusion of random effects (differences between questions and raters) significantly improves the explanatory power of the model.

The residual variance (σ^2^) is 3.29 and indicates the variability in the responses that is not explained by the model. The intergroup variance for the questions (τ00 ID_Q) is 5.22 and indicates significant variability in responses between different questions. The intraclass correlation coefficient (ICC) is 0.61, which means that about 61% of the total variability of the answers can be attributed to differences between the questions.

The chatbot responses to the same question show considerable variability, but this is slightly less than the variability between questions. The characteristics of the question (e.g. content, structure, context) had a significant impact on the differences in responses. The results suggest that while the independent variables alone have limited explanatory power, taking into account the specifics of individual questions and raters significantly improves the model’s ability to explain the variability of chatbot responses to exam questions.

The probability of obtaining a correct answer for ChatGPT-4 in English is OR=1.80, which is 0.64 when converted to probability. The probability of a correct answer for ChatGPT-4 in Polish is the same as for English. In contrast, the probability of a correct answer for Claude in Polish is 31% lower than for ChatGPT-4 in English.

### EMM

In the range for English, the probability ranged from 0.63 to 0.80, in the range for Polish from 0.48 to 0.77 (Table 7). For Polish, a non-linear relationship was observed with a decreasing tendency in the range up to Gemini and a substantial increase in probability for Claude. For both languages, the probability of getting a correct answer to an exam question is highest for Claude compared to ChatGPT-4 and Gemini. Overall, Claude performed better than the other chatbots in both languages, while ChatGPT4 performed equally well in both prompt languages. Gemini performed similarly well to ChatGPT4 in English, but showed a decrease in performance for Polish questions.

**Table 7.**
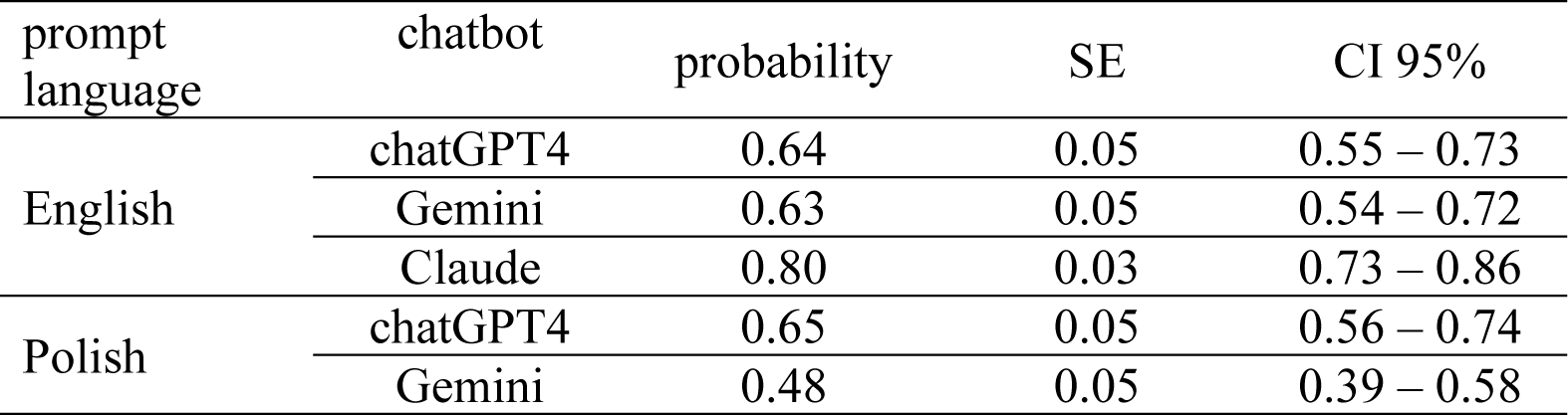
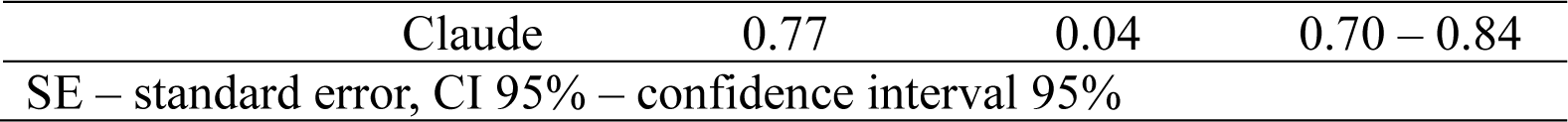
EMM - Estimated marginal means for the analysed variables (N = 594).

| prompt language | chatbot | probability | SE | CI 95% |
| --- | --- | --- | --- | --- |
| English | chatGPT4 | 0.64 | 0.05 | 0.55 – 0.73 |
|  | Gemini | 0.63 | 0.05 | 0.54 – 0.72 |
|  | Claude | 0.80 | 0.03 | 0.73 – 0.86 |
| Polish | chatGPT4 | 0.65 | 0.05 | 0.56 – 0.74 |
|  | Gemini | 0.48 | 0.05 | 0.39 – 0.58 |

|  |  |  |  |
| --- | --- | --- | --- |
| Claude | 0.77 | 0.04 | 0.70 – 0.84 |
| SE – standard error, CI 95% – confidence interval 95% |  |  |  |

### Analysis of Contrasts Between Chatbots Across Languages

The analysis of contrasts between chatbots across languages (Table 8, Figure 2) revealed significant differences in accuracy for both prompt languages (p < 0.001). The only exception was the comparison between ChatGPT-4 and Gemini in English, which showed no significant difference (p = 0.808).

Most differences showed a small effect size, with the exception of the comparison between Gemini and Claude in Polish, where a medium effect size was observed.

For English, ChatGPT-4 and Gemini showed similar effectiveness in predicting correct answers. The contrast analysis showed that both ChatGPT-4 and Gemini were less effective compared to Claude (p < 0.001). In the case of Polish, ChatGPT-achieved significantly better results than Gemini (p < 0.001); however, both chatbots were less effective than Claude (p < 0.001). The findings from these analyses led to the acceptance of the fifth hypothesis.

**Table 8.** Contrast analysis to investigate differences between chatbots across language categories.

| prompt language | contrast | OR | SE | CI 95% | p adjust |
| --- | --- | --- | --- | --- | --- |
| English | chatGPT4 / Gemini | 1.04 | 0.16 | 0.72 – 1.50 | 0.808 |
|  | chatGPT4 / Claude | 0.44 | 0.07 | 0.30 – 0.64 | **< 0.001** |
|  | Gemini / Claude | 0.43 | 0.07 | 0.29 – 0.62 | **< 0.001** |
| Polish | chatGPT4 / Gemini | 2.02 | 0.31 | 1.40 – 2.91 | **< 0.001** |
|  | chatGPT4 / Claude | 0.55 | 0.09 | 0.38 – 0.80 | **< 0.001** |
|  | Gemini / Claude | 0.27 | 0.04 | 0.19 – 0.40 | **< 0.001** |
| OR – odds ratio, SE – standard error, p adjust - p-value adjusted using the Holm correction |  |  |  |  |  |

**Figure 2:**
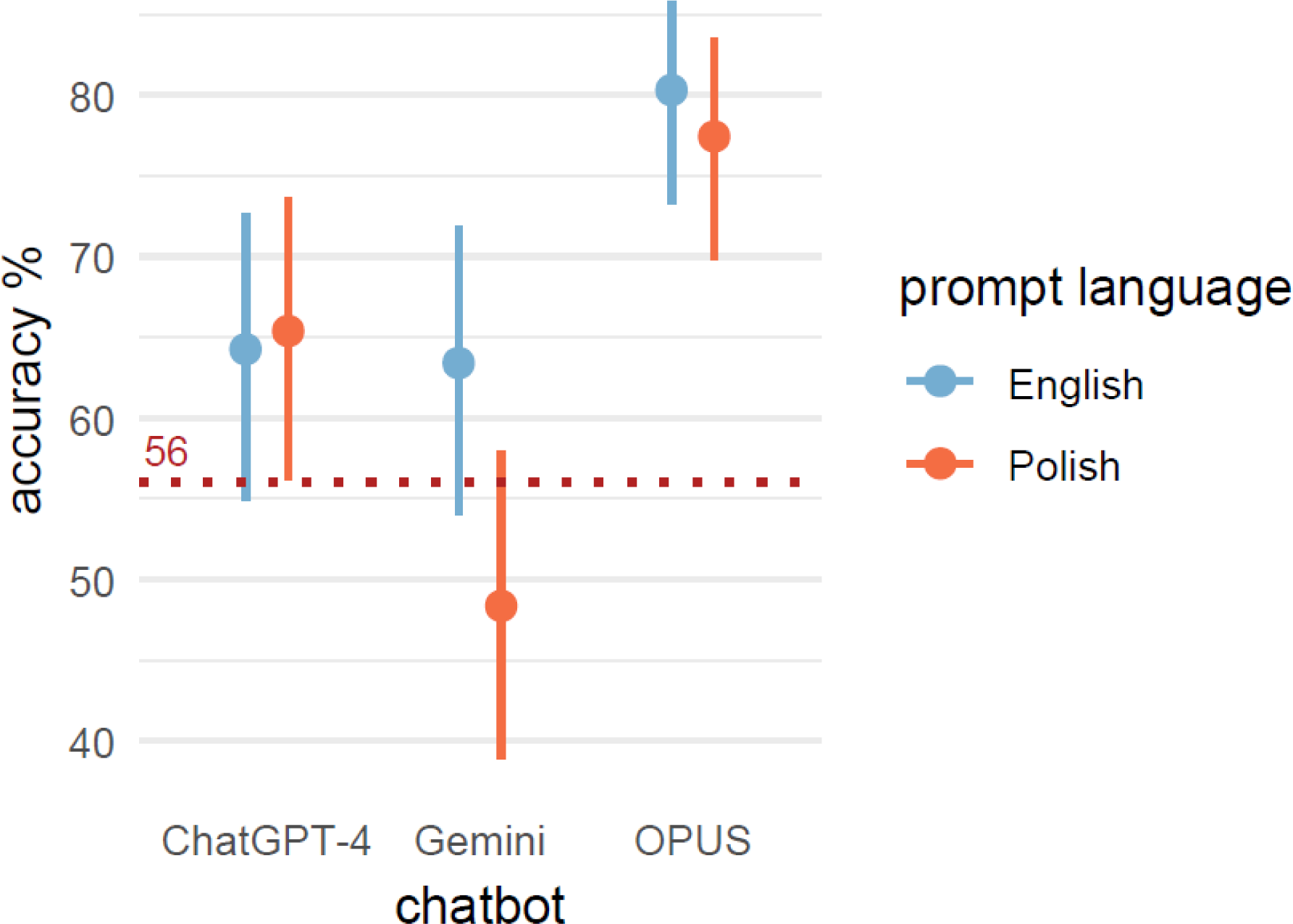
Differences in the probability of accuracy between chatbots in different language categories.

### Analysis of contrasts between chatbots within the prompt language

The Gemini model achieves significantly higher accuracy in English compared to Polish (Table 9, Figure 3). The odds ratio is 1.85, which means that the odds of the Gemini providing a correct answer are 84.5% higher in English than in Polish. This difference is statistically significant, with a small effect size.

For the ChatGPT-4 and Claude models, no statistically significant differences were observed in the accuracy of providing correct answers between English and Polish. The findings from this analysis support the acceptance of the sixth hypothesis.

**Table 9.**
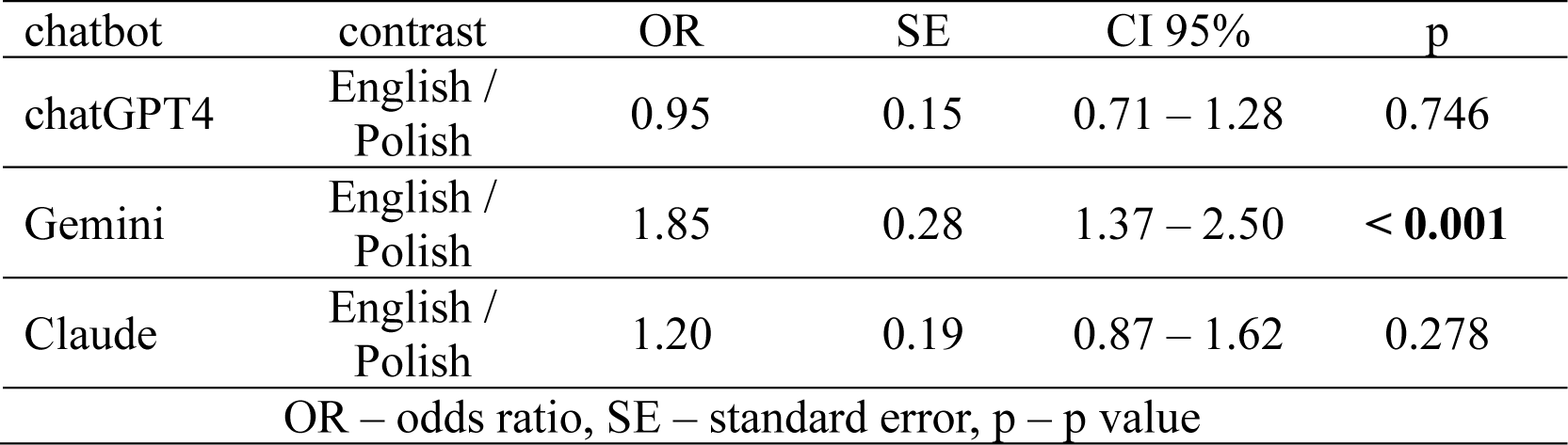
Intergroup differences between the languages for individual chatbots.

**Figure 3:**
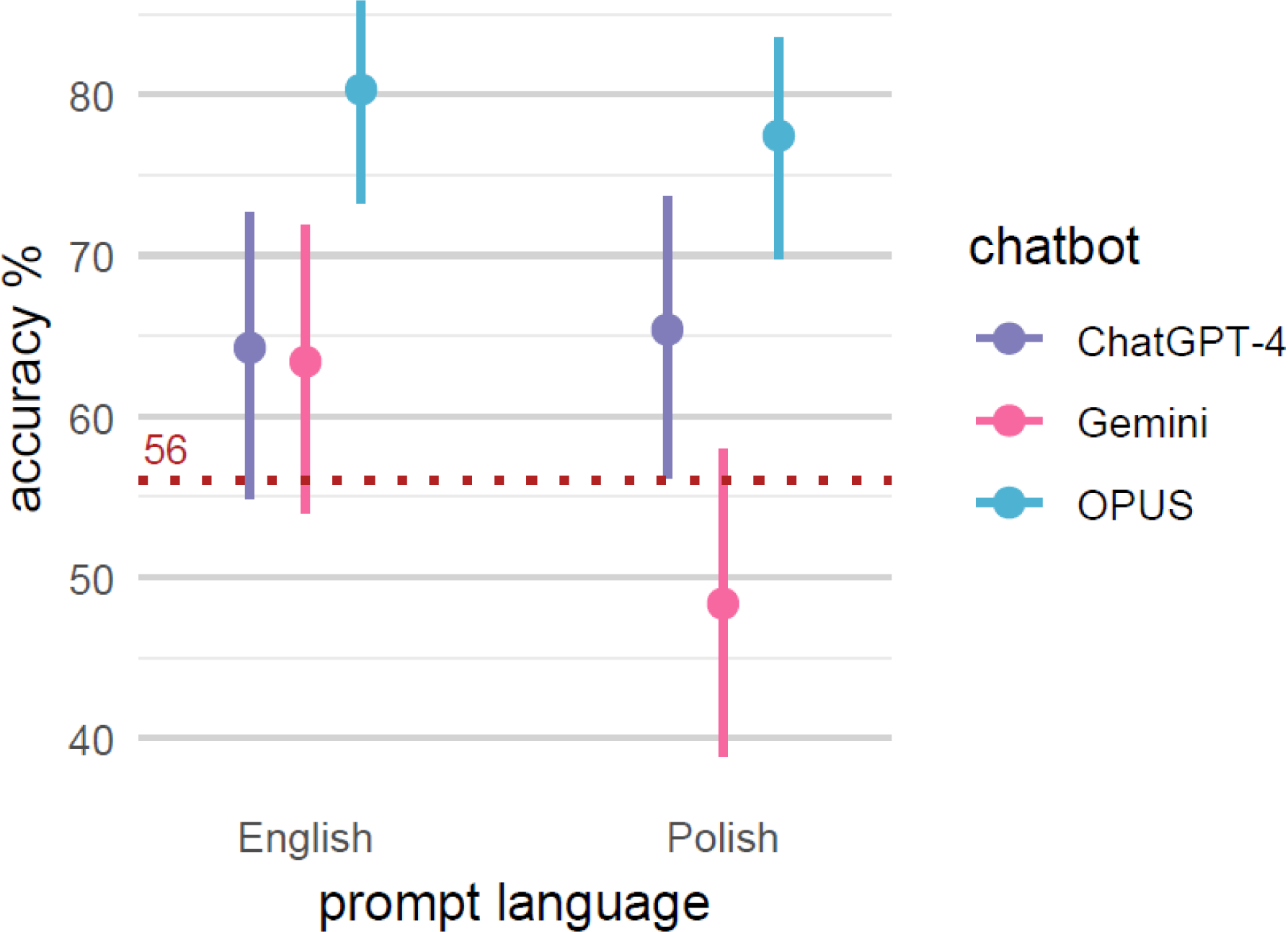
Differences in the probability of correct answers between languages within chatbots.

### Model 2

The reference categories for the chatbots are ChatGPT-4 responding to questions about conservative dentistry in English. The model was fitted on the basis of 3564 observations and 3 raters. The marginal R^2^ is 0.398. The random effects are as follows: σ^2^ = 3.29, τ00 for ID_Q = 5.19, τ11 ID_Q.languagepl = 1.24, ρ01 ID_Q = −0.15, ICC = 0.62. The model was fitted using 3564 observations and 3 raters.

The random effect τ11 represents the variance of random effects for the interaction between ‘ID_Q’ and ‘languagepl’. A variance of 1.24 indicates that there is some variability in how different questions are answered in Polish compared to English.

The random effect σ^2^ of 3.29 represents the residual variance or unexplained variability in the data after accounting for fixed and random effects. The τ00 for ‘ID_Q’ is 5.19, representing the variance of the random effects for the variable ‘ID_Q’, indicating differences between the questions.

The ρ01 value of −0.15 represents the correlation between the random effects for ‘ID_Q’ and ‘ID_Q.languagepl’, indicating a moderate negative correlation. This suggests that as the variance in one random effect increases, the variance in the other decreases to some extent.

The intraclass correlation coefficient (ICC) is 0.62. ICC measures the proportion of the total variance that is attributable to the grouping structure of the data (in this case, ‘ID_Q’). An ICC of 0.62 indicates that 62% of the variability in the probability of providing a correct answer can be attributed to differences between questions, rather than differences within questions. This suggests a substantial amount of clustering in the responses, which implies that the specific question being asked has a significant impact on the accuracy of the response.

The marginal R^2^ is 0.208, indicating that the fixed effects explain approximately 20.8% of the variability of the dependent variable, which is the probability of the chatbot providing a correct answer. The conditional R^2^ is 0.701, suggesting that the combined fixed and random effects explain 70.1% of the variability.

**Table 10.** EMM - Estimated marginal means for the analysed variables.

| question group | chatbot | English |  |  | Polish |  |  |
| --- | --- | --- | --- | --- | --- | --- | --- |
|  |  | probability | SE | CI 95% | probability | SE | CI 95% |
| conservative dentistry | chatGPT4 | 0.63 | 0.10 | 0.43 – 0.79 | 0.63 | 0.10 | 0.42 – 0.80 |
|  | Gemini | 0.58 | 0.10 | 0.38 – 0.75 | 0.39 | 0.10 | 0.21 – 0.60 |
|  | Claude | 0.83 | 0.06 | 0.68 – 0.92 | 0.85 | 0.06 | 0.71 – 0.93 |
| paediatric dentistry | chatGPT4 | 0.44 | 0.13 | 0.21 – 0.69 | 0.59 | 0.14 | 0.32 – 0.81 |
|  | Gemini | 0.53 | 0.13 | 0.28 – 0.76 | 0.51 | 0.14 | 0.26 – 0.76 |
|  | Claude | 0.77 | 0.10 | 0.53 – 0.91 | 0.87 | 0.07 | 0.69 – 0.96 |
| dental surgery | chatGPT4 | 0.59 | 0.13 | 0.33 – 0.81 | 0.32 | 0.13 | 0.13 – 0.59 |
|  | Gemini | 0.53 | 0.14 | 0.28 – 0.77 | 0.13 | 0.07 | 0.04 – 0.32 |
|  | Claude | 0.79 | 0.09 | 0.55 – 0.92 | 0.60 | 0.14 | 0.33 – 0.82 |
| prosthetic dentistry | chatGPT4 | 0.41 | 0.14 | 0.19 – 0.68 | 0.56 | 0.14 | 0.29 – 0.80 |
|  | Gemini | 0.53 | 0.14 | 0.27 – 0.77 | 0.54 | 0.14 | 0.27 – 0.78 |
|  | Claude | 0.43 | 0.14 | 0.20 – 0.70 | 0.33 | 0.13 | 0.14 – 0.61 |
| periodontology | chatGPT4 | 0.50 | 0.16 | 0.21 – 0.78 | 0.58 | 0.16 | 0.27 – 0.84 |
|  | Gemini | 0.60 | 0.16 | 0.29 – 0.84 | 0.17 | 0.10 | 0.05 – 0.44 |
|  | Claude | 0.69 | 0.14 | 0.38 – 0.89 | 0.57 | 0.16 | 0.27 – 0.84 |
| orthodontics | chatGPT4 | 0.37 | 0.15 | 0.14 – 0.67 | 0.66 | 0.15 | 0.35 – 0.88 |
|  | Gemini | 0.61 | 0.15 | 0.31 – 0.85 | 0.39 | 0.16 | 0.15 – 0.70 |
|  | Claude | 0.58 | 0.16 | 0.28 – 0.83 | 0.61 | 0.16 | 0.30 – 0.85 |
| integrated medicine | chatGPT4 | 0.98 | 0.01 | 0.94 – 1.00 | 0.94 | 0.03 | 0.85 – 0.98 |
|  | Gemini | 0.88 | 0.06 | 0.72 – 0.95 | 0.94 | 0.03 | 0.83 – 0.98 |
|  | Claude | 1.00 | 0.01 | 0.98 – 1.00 | 0.99 | 0.01 | 0.96 – 1.0 |

### Summary of chatbot performance in different specialties and languages

The lowest probability of providing a correct answer was 0.13 and was observed for Gemini in the specialty of oral surgery in Polish (Table 10). The highest probability was almost 1.0 for Claude in the specialty of integrated medicine, in both languages. The highest probabilities for all chatbots were observed in the specialty of integrated medicine for both languages. The lowest probabilities were observed for Gemini and ChatGPT-4 in Polish in the field of dental surgery.

### Analysis of contrasts between chatbots across question specialties and languages

#### English language

No statistically significant differences were detected among the chatbots in the specialties of prosthetic dentistry, periodontology, and orthodontics regarding the probability of accuracy to exam questions in English (Table 11, Figure 4).

In the specialty of integrated medicine, ChatGPT-4 exhibited over 7 times higher probability of accuracy compared to Gemini (p < 0.001). Gemini showed 96% lower probability of accuracy compared to Claude (p < 0.001). Both effect sizes are large.

#### Polish language

In the specialty of conservative dentistry, ChatGPT-4 showed over 7 times higher probability of accuracy compared to Gemini (p = 0.004, medium effect size). Gemini showed almost 90% lower probability of accuracy compared to Claude (p < 0.001, large effect size).

No statistically significant differences were detected among the chatbots in the specialties of prosthetic dentistry and orthodontics regarding the probability of accuracy to exam questions.

In the specialty of periodontology, ChatGPT-4 showed 6.76 times higher probability of providing a correct answer compared to Gemini (p = 0.002). Gemini showed 85% lower probability of providing a correct answer compared to Claude (p = 0.002). Both effect sizes are large.

Based on these results, the seventh hypothesis is supported.

**Table 11:**
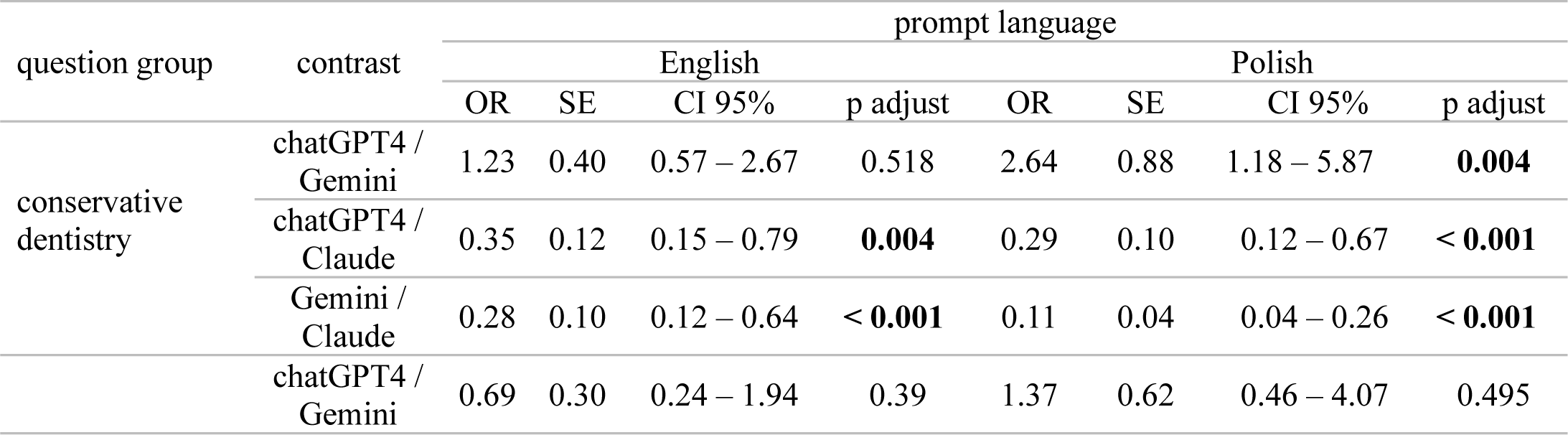
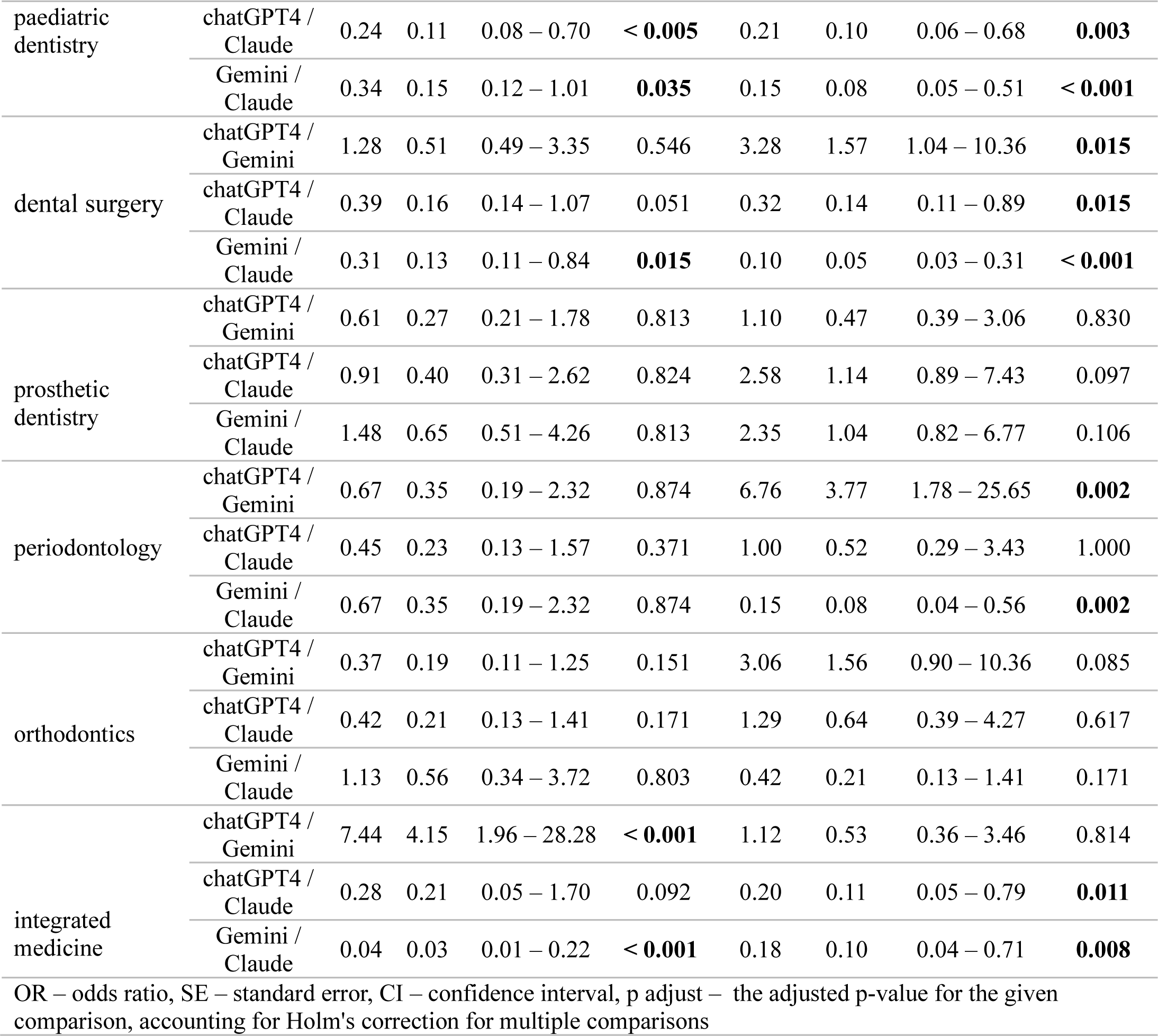
Analysis of accuracy differences across question specialties and prompt language categories between chatbots.

**Figure 4:**
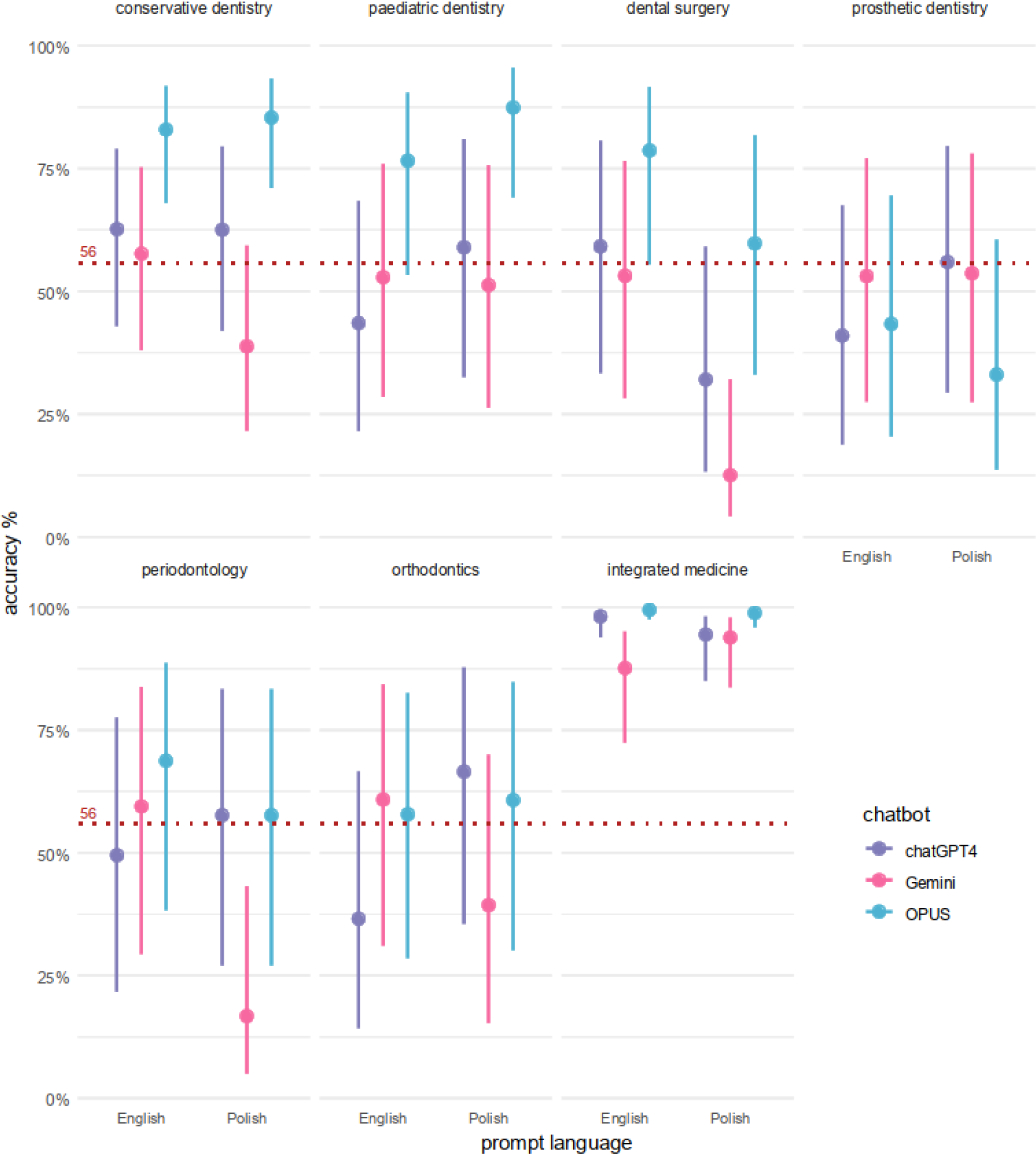
Analysis of differences between group within question specialties based on prompt language for chatbots.

### Model 1 Analysis (GLMER)

The analysis of GLMER Model 1 demonstrated that the probability of obtaining a correct answer by ChatGPT-4 in English was 0.64 (OR = 1.80), indicating its high efficacy in this language. However, the interaction between the Gemini chatbot and the Polish language significantly reduces the chances of getting a correct answer.

The random effects of Model 1 indicate substantial variability between question groups (ID_Q), suggesting variability in the likelihood of obtaining a correct answer depending on the question group. Approximately 61% of the total variance in responses can be attributed to differences between question groups. Model 1 as a whole (including random effects) explains 62,4% of the variance in responses, while fixed effects alone explain only 2.7% of the variance.

### Model 2 Analysis

The analysis of Model 2, which compared chatbots within question groups, showed that the chatbot Claude achieved the highest probability of accuracy for all question groups except the area of prosthetic dentistry compared to ChatGPT-4 and Gemini. In addition, the probability of a correct answer to questions in the field of integrated medicine is higher than in the field of dentistry for all chatbots in both prompt languages.

### Performance differences

The differences in performance between chatbots in English and Polish in some areas (e.g. integrated medicine) indicate that the language of the prompt influences the probability of accuracy.

## Discussion

In this study, chatbots showed significant consistency in their responses between English and Polish. The results of Cohen’s Kappa analysis show that Claude had the highest consistency in accuracy regardless of the prompt language. These results indicate that Claude gave the most consistent answers to the same dental examination questions regardless of the number of measurements. All chatbots showed satisfactory inter-rater reliability, emphasising their potential to generate consistent answers in dental and medical contexts. Remarkably, Gemini showed the lowest agreement in answers among the analysed chatbots, with a surprisingly lower consistency in English prompts compared to Polish ones. It is important to note that this data does not reflect the correctness of the chatbots’ answers compared to the answer key. These results formed the basis for the inclusion of the language variable in both generalised linear mixed models (glmer).

Model 1, which includes both fixed and random effects, explains 62,4% of the variance in responses, while fixed effects alone explain only 2.7% of the variance. This suggests that the characteristics of the questions (e.g. content, structure, context) significantly influence the differences in chatbot responses.

Model 2 showed that the probability of a correct answer to questions in the field of integrated medicine is higher for all chatbots than in the field of dentistry, regardless of the language of the prompt. This indicates that Claude has better knowledge and skills in the field of integrated medicine than in dentistry.

In recent years, there has been growing interest in the ability of chatbots to pass medical exams. Increasingly, researchers are investigating the potential of chatbots to enhance the education of medical students and professionals. Analysing the accuracy of chatbots can help improve these tools and enable their more effective use in the educational process. It can also help users to select the chatbot that works best in a particular area, which could lead to better support in medical education and practise.

A recent study of Chau et al. examined the performance of ChatGPT 4.0, in dental licensing exams. Researchers posed 1,461 multiple-choice questions from dental licensing exams in the US and UK to two versions of chatGPT - chatGPT 3.5 and chatGPT 4.0.

ChatGPT 4.0 passed both tests, answering 80.7% of the questions correctly in the US dental licensing exam and 62.7% correctly in the UK dental licensing exam^19^. In this comparative analysis, in the Polish dental licensing exam, ChatGPT 4.0 correctly answered 58% of the questions in English and 61% of the questions in Polish, thereby achieving the required score.

A study of Ahmad et al. compared the performance of ChatGPT-4o, Claude 3 OPUS, and Gemini Advanced on the 2024 periodontology in-service examination. ChatGPT-4o achieved the highest accuracy in all domains, ranging from 85.7% to 100%, significantly outperforming Claude 3 OPUS and Gemini Advanced^20^. Our study showed statistically significant differences between chatbots in periodontology in terms of the probability of accuracy only for exam questions in Polish. ChatGPT-4 showed 6.76 times higher probability of providing a correct answer compared to Gemini. Gemini showed 85% lower probability of providing a correct answer compared to Claude.

There are also some recent studies on medical licensing exams.

Kung et al. demonstrated in their research that ChatGPT exhibited moderate performance on the USMLE exam, with accuracy rates varying between 45.4% and 75.0% based on question type, approaching but not consistently achieving passing scores.^21^.

Weng et al. evaluated the performance of ChatGPT on Taiwan’s Family Medicine Board Exam. Surprisingly, ChatGPT answered 52 out of 125 questions correctly, which corresponds to an accuracy rate of 41.6%, which was insufficient to pass the exam^22^.

A study by D’Anna et al. evaluated the performance of ChatGPT 4 and Google Bard on European Society of Neuroradiology (ESNR) course exams. ChatGPT 4 showed an overall accuracy of 70%, clearly outperforming Google Bard. ChatGPT 4 passed all four multiple-choice exams (MCEs) from the ESNR courses, while Google Bard did not^23^.

Several studies have evaluated the performance of chatbots, particularly ChatGPT, in Polish medical exams. ChatGPT Fails Polish Board Certification Examination in Internal Medicine. ChatGPT’s performance on the PES in internal medicine was challenging, with an average success rate of 49.4%, falling short of the 60% passing threshold.^1^

In another study, ChatGPT-3.5 was reported to have passed the final medical examination in Poland. The performance was satisfactory, but still worse than that of human doctors, as they answered 53.4% to 64.9% of the questions correctly. In 8 out of 11 exam sessions, ChatGPT achieved the score required to pass the examination (60%).^4^.

These findings highlight the potential of AI tools like ChatGPT 4.0 to assist in medical education by providing accurate and reliable responses in multiple languages and contexts.

### Limitations of the study

This study focussed on investigating the accuracy of three chatbots: ChatGPT4, Gemini and Claude. The analysis focused on specific versions of these chatbots, which may affect the generalisability of the results to other models. Moreover, the results may vary depending on the prompt language. Our study focused on Polish and English, which may limit the generalisability of the results to other languages. The investigation was limited to a single, recent examination paper. Whilst this approach provides current insights, it may not fully represent the performance of chatbots on a wider range of dental topics or question types.

## Conclusions

### 1. Self-assessment and effectiveness

The results suggest that chatbots have some ability to recognise the relative difficulty of questions, but the accuracy of this assessment is limited.

### 2. Self-evaluation and actual performance

To some extent chatbots can estimate their answers reasonably correctly. However, caution is required when interpreting the results. The small effect size indicates that the chatbots’ self-assessment may not be a completely reliable indicator of their actual effectiveness.

### 3. Linguistic consistency

Chatbot responses to the same questions may vary depending on the language of the prompt. This linguistic variability should be considered when using chatbots in multilingual contexts or for tasks requiring consistency between languages.

### 4. Measurement consistency

The results suggest that chatbot responses to the same questions are not completely predictable or repeatable. Differences in consistency can result from various factors: model updates between measurements, built-in randomness in generating responses, differences in interpreting the context of questions at different points in time.

### 5. Accuracy in different prompt languages

Chatbots differ in the accuracy of providing correct answers for individual prompt languages. The most significant differences occur between Gemini and Claude in English and between ChatGPT-4 and Gemini in Polish. Claude has the highest accuracy in both languages. The performance differences are more pronounced in Polish than in English, especially when you compare Gemini with other chatbots.

### 6. Accuracy across all languages

Only Gemini differs significantly in accuracy of correct answers between Polish and English prompts. The accuracy of Gemini is significantly (by 31.25%) higher in English than in Polish. For the other chatbots, no differences in accuracy were found between the languages, with ChatGPT-4 and Claude answering the exam questions equally well in both English and Polish.

### 7. Specialty-specific accuracy

Chatbot accuracy is highest in the integrated medicine domain, indicating higher accuracy for general medicine questions than dental questions. The Claude chatbot showed the highest effectiveness in providing correct answers regardless of the language (English or Polish) in which the exam questions were asked, especially in the areas of conservative dentistry, paediatric dentistry, dental surgery and integrated medicine. ChatGPT-4 showed greater effectiveness than Gemini in generating correct answers to exam questions in some fields, such as dental surgery and periodontology. No significant differences were found between the chatbots in the areas of prosthetic dentistry and orthodontics, suggesting similar effectiveness of all three models in these areas. The choice of an appropriate chatbot may depend on the language of the prompt and the specific area of dentistry or integrated medicine to which the question relates.

Our results demonstrate that Claude achieved the highest accuracy in all areas analysed and outperformed other chatbots. This suggests that Claude has significant potential to support the medical education of dental students.

This study showed that the performance of chatbots varied depending on the prompt language and the specific field. For instance, ChatGPT-4 showed better performance than Gemini in some areas, especially in Polish. This highlights the importance of considering language and specialty when selecting a chatbot for educational purposes.

While chatbots show promise in supporting medical education, their use requires caution and a deliberate approach to avoid errors and misinformation. Further research is essential to better evaluate the potential of chatbots in medical education. Developing critical thinking skills in students is crucial to enable them to effectively evaluate the information generated by chatbots.

## Data Availability

data will be awailable at Mendeley repository

## References

1. Wójcik, S. et al. Reshaping medical education: Performance of ChatGPT on a PES medical examination. Cardiol. J. (2023).

2. Lewandowski, M., Łukowicz, P., Świetlik, D. & Barańska-Rybak, W. An original study of ChatGPT-3.5 and ChatGPT-4 dermatological knowledge level based on the dermatology specialty certificate examinations. Clin. Exp. Dermatol. llad255 (2023).

3. Gilson, A., et al. How does ChatGPT perform on the United States Medical Licensing Examination (USMLE)? The implications of large language models for medical education and knowledge assessment. JMIR Med. Educ. 9, e45312 (2023).

4. Suwała, S. et al. ChatGPT-3.5 passes Poland’s medical final examination—Is it possible for ChatGPT to become a doctor in Poland? SAGE Open Med. 12, 20503121241257777 (2024).

5. Funder, D. C. & Ozer, D. J. Evaluating effect size in psychological research: Sense and nonsense. Adv. Methods Pract. Psychol. Sci. 2, 156–168 (2019).

6. Cohen, J. Statistical Power Analysis for the Behavioral Sciences. (New York: Routledge, 1988).

7. McHugh, M. L. Interrater reliability: the kappa statistic. *Biochem*. Medica 22, 276–282 (2012).

8. Fleiss, J.L., Levin, B., & Paik, M.C. Statistical methods for rates and proportions. (2003).

9. Powell, M. J. The BOBYQA algorithm for bound constrained optimization without derivatives. Camb. NA Rep. NA200906 Univ. Camb. Camb. **26**, 26–46 (2009).

10. Wickham, H. & Bryan, J. readxl: Read Excel Files. R package version 1.3. 1. (2019).

11. Wickham H, François R, Henry L, Müller K, Vaughan D. dplyr: A Grammar of Data Manipulation. R package version 1.1.4,. (2023).

12. Ben-Shachar, M. S., Lüdecke, D. & Makowski, D. effectsize: Estimation of effect size indices and standardized parameters. J. Open Source Softw. 5, 2815 (2020).

13. Bates, D., Mächler, M., Bolker, B. & Walker, S. Fitting Linear Mixed-Effects Models Using lme4. J. Stat. Softw. 67, 1–48 (2015).

14. Lüdecke, M. D. Package ‘sjPlot’. (2024).

15. Gamer, M., Lemon, J., Gamer, M. M., Robinson, A. & Kendall’s, W. Package ‘irr’. Var. Coeff. Interrater Reliab. Agreem. 22, 1–32 (2012).

16. Wickham, H. Tidyr: Tidy Messy Data: R Package Version 1.3.1 2024. URL HttpsCRAN R-Proj. Orgpackage Tidyr (2024).

17. Lüdecke, D. et al. see: An R package for visualizing statistical models. J. Open Source Softw. 6, 3393 (2021).

18. Wickham, H. & Wickham, H. Data Analysis. (Springer, 2016).

19. Chau, R. C. W. et al. Performance of generative artificial intelligence in dental licensing examinations. Int. Dent. J. 74, 616–621 (2024).

20. Ahmad, B., Saleh, K., Alharbi, S., Alqaderi, H. & Jeong, Y. N. Artificial Intelligence in Periodontology: Performance Evaluation of ChatGPT, Claude, and Gemini on the In-service Examination. medRxiv 2024.05.29.24308155 (2024) doi:10.1101/2024.05.29.24308155.

21. Kung, T. H. et al. Performance of ChatGPT on USMLE: potential for AI-assisted medical education using large language models. *PLoS Digit*. Health 2, e0000198 (2023).

22. Weng, T.-L., Wang, Y.-M., Chang, S., Chen, T.-J. & Hwang, S.-J. ChatGPT failed Taiwan’s family medicine board exam. J. Chin. Med. Assoc. 86, 762–766 (2023).

23. D’Anna, G., Van Cauter, S., Thurnher, M., Van Goethem, J. & Haller, S. Can large language models pass official high-grade exams of the European Society of Neuroradiology courses? A direct comparison between OpenAI chatGPT 3.5, OpenAI GPT4 and Google Bard. Neuroradiology 1–6 (2024).

